# Beyond Motor Fluctuations: Understanding the Clinical Correlates of OFF burden in Parkinson’s Disease

**DOI:** 10.64898/2026.04.04.26350175

**Authors:** David Ledingham, Sahana Sathyanarayana, Robyn Iredale, Charlotte B. Stewart, Victoria Foster, Debra Galley, Mark R Baker, Nicola Pavese

## Abstract

**Background:** Historically, OFF burden in Parkinson’s disease has been primarily attributed to motor features. Recent studies highlight that non-motor symptoms, and the predictability of OFF episodes also drive functional impairment, yet they are rarely measured in clinical practice.

**Objective:** To identify which clinical features are most closely associated with OFF time and OFF impact, and to quantify the added explanatory value of temporal predictability, non-motor, and behavioural domains beyond a core motor model.

**Methods:** We analysed 1,252 OFF-only visits from 430 PPMI participants. Outcomes were MDS-UPDRS IV 4.3 (OFF time) and 4.4 (OFF impact). Linear mixed-effects models with a participant random intercept were fitted. The core motor model included OFF-state motor severity, freezing, tremor, levodopa responsiveness, and dyskinesia, plus covariates. Predictability (IV 4.5), non-motor (mood, fatigue/sleep, autonomic/GI), and behavioural (impulse-control behaviours) domains were then added to assess added influence beyond motor. Analyses were stratified by time since diagnosis (Pooled; ≤ 4 y; ≥ 6 y).

**Results:** Clinical features explained more variance in OFF impact than OFF time (25.9% vs 8.1%). OFF time was primarily linked to OFF-state motor severity/freezing, with levodopa responsiveness important early. For OFF impact, predictability produced the largest increment in marginal R² beyond the core motor model (pooled and Late). Within the core motor model, tremor was the largest contributor to OFF impact.

**Conclusions:** Predictability is a prominent correlate of OFF impact. Asking about predictability may help tailor therapy, from timing optimisation to on-demand rescue for unpredictable episodes.

**Plain Language Summary:** People with Parkinson’s disease often experience “OFF periods,” when their usual medication stops working and symptoms return. These episodes can make everyday activities difficult. Traditionally, OFF periods have been measured by how much time they last, but patients often say that unpredictability, when OFF episodes happen without warning, is even more disruptive.

Our study looked at data from over 1,200 clinic visits in a large international research project. We examined two aspects of OFF burden:

1. OFF time – how much of the day is spent in an OFF state.
2. OFF impact – how much OFF episodes interfere with daily life.

We correlated these with motor symptoms (such as tremor and freezing), non-motor symptoms (such as anxiety and fatigue), and a measure of predictability (how regular or irregular OFF episodes are).

We found that OFF impact was strongly linked to predictability. However, this does not mean unpredictability alone makes OFF worse, it may reflect a different type of OFF episode. Predictable “wearing-off” usually occurs gradually as medication wears off, while “on–off fluctuations” can happen suddenly and are often more severe. Our findings suggest that patients who experience these abrupt changes report greater disruption to daily life.

Why does this matter? Asking patients whether they can predict their OFF episodes may help doctors choose the right treatment. Predictable wearing-off can often be managed by adjusting medication timing or adding long-acting drugs. On–off fluctuations may need fast-acting rescue treatments. In some cases, frequent unpredictable OFF episodes may signal the need to consider advanced options like infusion therapies or deep brain stimulation earlier in care.

Our findings suggest that predictability should be part of routine assessment, alongside motor symptoms. Future research should explore whether improving predictability or targeting these more severe fluctuations can reduce the impact of OFF periods.

## Introduction

OFF periods in Parkinson’s disease (PD) have traditionally been understood as fluctuations in motor features arising from disease progression and the pharmacokinetics of dopaminergic therapy(1). Longitudinal and trial data link the development of OFF/motor complications to dopaminergic exposure, levodopa responsiveness, younger age, female sex, disease duration, and motor severity (2–4).

Beyond these motor- and medication-centric risk factors, non-motor features (i.e. gastrointestinal dysfunction, fatigue/sleep, mood, and impulse-control behaviours) shape OFF burden by influencing levodopa handling (e.g., GI absorption), day-to-day resilience, and the perceived impact of OFF episodes. Prominent OFF periods may, in turn, amplify distress and non-motor symptoms(4, 5). Prospective cohort data indicate that baseline non-motor burden, particularly low mood and anxiety, increases subsequent risk of motor complications(4). Patient-reported studies further show that the OFF experience is multidimensional: fluctuations in emotional distress, fatigue, autonomic symptoms, and the unpredictability of off periods are sometimes more disruptive than fluctuations in motor features(6–8). Yet these dimensions are not consistently captured in routine clinical assessment.

Despite detailed qualitative descriptions of OFF experiences, no study has attempted to rank the relative contributions of different clinical domains to OFF burden within a single longitudinal framework(6, 7). Most studies treat OFF as a binary phenomenon (present/absent) and focus on OFF time rather than its functional impact. Using longitudinal data from the Parkinson’s Progression Markers Initiative (PPMI), we set out to identify which clinical features are most clearly associated with two complementary outcomes, OFF time and OFF impact, and to quantify their relative contributions across the disease course. We adopted a staged quantitative framework: first establishing a core motor model (OFF-state motor severity, freezing, tremor, levodopa responsiveness, and dyskinesia with covariates), then testing whether temporal predictability, non-motor symptoms and impulse-control behaviours add explanatory value beyond that motor baseline.

## Methods

### Study Design and Participants

We used longitudinal data from the PPMI study, a multicentre observational study designed to identify PD progression markers (ClinicalTrials.gov: NCT01141023)(9). All participants provided informed consent, and ethics approval was obtained at each site.

We included participants with sporadic PD who had initiated dopaminergic therapy and contributed at least one visit with a non-zero score for MDS-UPDRS Part IV item 4.3 (OFF time). Item 4.4 (OFF impact) could be zero. Visits wherein both items scored zero were excluded, as the aim was to examine correlates of OFF burden rather than predictors of OFF onset. All visits after initiation of device-aided therapy (e.g., deep brain stimulation or infusion therapies) were excluded.

### Outcomes

OFF burden was assessed using two complementary MDS-UPDRS Part IV items: item 4.3 (OFF time), which captures the proportion of the waking day spent in OFF, and item 4.4 (OFF impact), which reflects the functional impact of OFF episodes. Both outcomes were analysed as continuous (for standardised effects and ΔR²), assuming the ordinal categories approximate equal spacing, a convention applied in longitudinal MDS-UPDRS research(10) and supported by evidence that parametric methods perform well for ordinal scales at large sample sizes(11). Sensitivity analyses using ordinal cumulative link mixed models (CLMM) were performed to confirm consistency.

### Predictor selection and mapping

Predictors domains were selected based on the clinical literature identifying factors that influence the severity and impact of OFF periods, including motor features and treatment response, non-motor symptoms that modify perceived burden, temporal characteristics, and behavioural complications. Each domain was mapped to the best available validated PPMI measure, as detailed in Supplementary Tables 1A and 1B.

Motor severity during OFF was assessed using the MDS-UPDRS Part III OFF total score. To capture fluctuating symptoms, we included patient-reported freezing (Part II item 2.13) and tremor (Part II item 2.10). These symptoms are consistently rated among the most bothersome during OFF periods in patient surveys(6, 7). We did not include PIGD vs tremor-dominant subtypes, as PIGD scoring overlaps with Part III items and subtype classification varies with medication state and disease duration. Our aim was to focus on specific symptoms that patients identify as impactful, rather than composite phenotypes. Levodopa responsiveness was calculated as the percentage improvement in Part III scores from OFF to ON when paired examinations were available. Dyskinesia burden was assessed using the mean of Part IV items 4.1 and 4.2 (time and functional impact of dyskinesia respectively).

Non-motor symptoms included anxiety (STAI-State and Trait), depression (GDS-15), fatigue and sleepiness (Epworth Sleepiness Scale), autonomic symptoms (SCOPA-AUT total excluding Gastrointestinal (GI) items), and gastrointestinal symptoms (SCOPA-AUT GI domain).

Non-motor domains were measured using standard instruments at each visit but not timed to OFF periods. Associations should therefore be interpreted as reflecting overall burden rather than OFF-specific symptom change.

Predictability of fluctuations was assessed using MDS-UPDRS Part IV item 4.5 and treated as a temporal feature. Impulse-control behaviours were assessed using the Questionnaire for Impulsive-Compulsive Disorders in Parkinson’s Disease (QUIP)(12). These were included as they can modify the experience of OFF burden, for example through dose escalation and dopamine withdrawal effects, and destabilise treatment regimens.

Covariates included sex, age at onset, and disease duration to adjust for demographic and duration-related differences. Levodopa equivalent daily dose (LEDD) was summarised as the person-mean across OFF-only visits and treated as a stable exposure covariate to reduce bidirectional bias. This approach acknowledges that higher doses may reflect greater disease severity but may also contribute to motor complications and OFF fluctuations. Time-varying predictors were decomposed into within-person (visit-level deviation) and between-person (person-mean) components. This allowed us to differentiate longitudinal change from stable differences across individuals and improves interpretability of mixed-effects models.

### Statistical Analysis

We used linear mixed-effects models (LMMs) with a random intercept for participant to analyse OFF time (MDS-UPDRS IV 4.3) and OFF impact (IV 4.4). Models were fit in R (lme4/lmerTest) by restricted maximum likelihood (REML). Outcomes and predictors were z-standardised. Time-varying variables were split into within-person (visit-level change) and between-person (person-mean) components.

First, we fitted the core motor model, defined here as the prespecified motor feature set with covariates retained throughout: MDS-UPDRS III OFF total; patient-reported freezing (II-2.13) and tremor (II-2.10); levodopa responsiveness (% OFF→ON change in III where paired exams were available); dyskinesia burden (mean IV 4.1 time + IV 4.2 impact); covariates were sex, age at onset, disease duration, and person-mean LEDD (exposure). Model fit was summarised by marginal R² (R²m; fixed effects) and conditional R² (R²c; fixed + random). We also report the percentage change in random-intercept variance when models were expanded.

Second, we evaluated added value beyond the core motor baseline by adding domain blocks: Predictability (IV 4.5), Non-motor (anxiety, depression, fatigue/sleep, autonomic, gastrointestinal), and Behavioural (impulse-control behaviours). For domain tests, ΔR² was defined as the increase in R²m when the domain block was added (an add-one approach) to the core motor model baseline. Because blocks are correlated, ΔR² values are not additive; in figures, the stacks are for visual guidance: they show the improvement when each domain is added to the core motor model. Because the domains overlap, the segments shouldn’t be summed as if they were separate shares.

Third, we separated endpoint overlap using conditioned residual analyses. For each OFF outcome, we fitted the core motor model (with covariates) and took fixed-effects residuals (the portion not explained by motor + covariates). We then modelled these residuals with axis domains and quantified axis-level contributions using drop-one ΔR². This approach isolates associations with the portion of OFF burden not explained by motor features and complements the incremental ΔR² analyses. While the incremental models summarise associations across all visits and quantify which domains contribute most to OFF burden at the cohort level, mismatch residual modelling focuses on what explains differences in perceived burden among patients with similar motor severity. In a sensitivity version, the base model also included the alternate OFF endpoint before residuals were taken (for example, OFF impact residualised on motor and OFF time), so ΔR² reflects added value beyond motor and the other OFF measure.

Analyses were run pooled and duration-stratified by time since diagnosis on a visit basis (Early ≤ 4 years; Late ≥ 6 years). Grouping was visit-based, allowing participants to contribute to both groups as their disease progressed; visits with duration 4–<6 years were excluded from group-specific analyses but included in pooled analyses. These thresholds were chosen pragmatically to maximize contrast between an early population and a more established population, as previous work in the PPMI cohort has shown that OFF periods affect ∼35% of patients by 4–6 years and ∼59% by 8–10 years(3).

Finally, Given the ordinal (0–4) scoring of IV 4.3/4.4, we ran cumulative-link mixed models (CLMM) as a sensitivity analysis that respects the ordinal scale. CLMM analyses were restricted to the Late (≥ 6 years) group where greater heterogeneity was expected. We report Cox–Snell and Nagelkerke pseudo R² and odds ratios per 1 SD.

Models were fit on complete cases per specification; sample sizes vary by domain availability, responsiveness availability, and disease-duration filters.

## Results

### Participant characteristics

Analyses included 1,252 OFF-only visits from 430 participants in the PPMI cohort. Of these, 213 participants contributed to the ≤4-year group (265 visits), and 233 participants to the ≥6-year group (760 visits). Participants in the late group were older at visit and had higher LEDD and greater OFF-state motor severity. Levodopa responsiveness was available for 67.5% of visits. OFF time was mild across groups, while OFF impact was more frequent and showed greater variability, particularly in the ≥6-year group. Any OFF impact was reported in 81.8% of visits in the ≥6-year group compared with 59.2% in the ≤4-year group. Non-motor domains also differed by group, with higher anxiety and autonomic burden in the ≥6-year group, and modest increases in sleepiness and gastrointestinal symptoms. Predictability scores were low (median 1), but scores ≥2 were more frequent in the ≥6-year group (22.5% vs 16.2%).

Table 1 summarizes core descriptors; extended non-motor distributions are provided in Supplementary Table S2 and 3.

**Table 1.** Baseline characteristics of OFF-only visits by disease duration. Values are presented as mean ± SD or median [IQR] where appropriate. Acronyms: LEDD, levodopa equivalent daily dose; MDS-UPDRS, Movement Disorder Society-Unified Parkinson’s Disease Rating Scale; OFF, medication “off” state; ON, medication “on” state; IQR, interquartile range.

| Variable | Pooled | Early ($\leq 4$ y) | Late ($\geq 6$ y) |
| --- | --- | --- | --- |
| Visits (n) | 1,252 | 265 | 760 |
| Participants (n) | 430 | 213 | 233 |
| Sex = male (%) | 66.7 | 58.9 | 69.3 |
| Age at visit (y), mean ± SD | 67.3 ± 9.6 | 64.4 ± 9.9 | 68.8 ± 9.0 |
| Age at onset (y), mean ± SD | 60.1 ± 9.3 | 61.8 ± 9.9 | 59.3 ± 8.7 |
| Disease duration (y), mean ± SD | 7.2 ± 3.5 | 2.6 ± 0.8 | 9.5 ± 2.4 |
| LEDD (mg/day), mean ± SD | 814 ± 521 | 499 ± 339 | 963 ± 535 |
| MDS UPDRS-III OFF, mean ± SD | 35.1 ± 14.3 | 30.3 ± 12.9 | 38.0 ± 15.0 |
| MDS UPDRS-III ON, mean ± SD | 24.1 ± 13.2 | 21.1 ± 11.8 | 26.0 ± 13.7 |
| Responsiveness available (%) | 67.5 | 74.3 | 63.7 |
| OFF time (4.3), median [IQR] | 1 [1–2] | 1 [1–2] | 1 [1–2] |
| 4.3 Time in OFF (1–4) distribution | 1: 78.0%;<br>2: 17.2%;<br>3: 3.0%;<br>4: 1.8% | 1: 78.5%;<br>2: 12.8%;<br>3: 3.4%;<br>4: 5.3% | 1: 76.8%;<br>2: 19.2%;<br>3: 3.3%;<br>4: 0.7% |
| OFF impact (4.4), median [IQR] | 1 [0–2] | 1 [0–1] | 1 [1–3] |
| Any OFF impact (%) | 75.3 | 59.2 | 81.8 |
| 4.4 OFF impact (0–4) distribution | 0: 24.7%;<br>1: 43.2%;<br>2: 15.6%;<br>3: 15.1%;<br>4: 1.4% | 0: 40.8%;<br>1: 45.7%;<br>2: 9.8%;<br>3: 3.4%;<br>4: 0.4% | 0: 18.2%;<br>1: 39.6%;<br>2: 19.1%;<br>3: 21.1%;<br>4: 2.1% |

### Core motor analysis

#### Linear Mixed-Effects Models

##### Model fit

The full mixed-effects models explained a substantial proportion of variance, with random intercepts accounting for most of the conditional R². Fixed effects explained more variance for OFF impact than OFF time across all strata. In pooled analyses, marginal R² was 8.1% for OFF time and 25.9% for OFF impact (conditional R²: 39.6% and 42.3%, respectively). Disease duration-specific models showed higher marginal R² for OFF time in the ≤4-year group (21.4%) than in the ≥6-year group (11.3%), whereas OFF impact remained stable (26.9% vs 20.8%). See Table 2.

**Table 2.** Model fit for mixed-effects models of OFF burden. Marginal R² represents variance explained by fixed effects; conditional R² represents variance explained by fixed and random effects combined. Higher conditional R² values indicate substantial participant-level variance captured by random intercepts. Acronyms: OFF, medication “off” state; R², coefficient of determination.

| Group | Outcome | Marginal R<sup>2</sup> (%) | Conditional R<sup>2</sup> (%) |
| --- | --- | --- | --- |
| Pooled | OFF time (4.3) | 8.1 | 39.6 |
| Pooled | OFF impact (4.4) | 25.9 | 42.3 |
| Early ($\leq 4$ y) | OFF time (4.3) | 21.4 | 65.4 |
| Early ($\leq 4$ y) | OFF impact (4.4) | 26.9 | 44.1 |
| Late ($\geq 6$ y) | OFF time (4.3) | 11.3 | 44 |
| Late ($\geq 6$ y) | OFF impact (4.4) | 20.8 | 38.6 |

##### Pooled analyses

For OFF time (item 4.3), within-person motor severity (MDS UPDRS III OFF score) showed the strongest association (β = 0.123, p < 0.001), with smaller effects for freezing (β = 0.063, p = 0.076) and tremor (β = 0.075, p = 0.019). Between patients, greater treatment responsiveness correlated with less OFF time (β = −0.131, p = 0.009).

For OFF impact (item 4.4), within-person associations were observed for motor severity (β = 0.125, p < 0.001), freezing (β = 0.093, p = 0.004), and dyskinesia (β = 0.091, p = 0.004). Between-person associations included freezing (β = 0.123, p = 0.006), tremor (β = 0.138, p < 0.001), and dyskinesia (β = 0.099, p = 0.019), along with covariate associations for LEDD (β = 0.140, p < 0.001) and disease duration (β = 0.281, p < 1e−12).

(See Table 3 for full estimates.)

**Table 3.** Fixed-effect estimates for mechanistic predictors and covariates in mixed-effects models of OFF burden. Values represent standardized coefficients (β) for within-person (“Across visits”) and between-person effects. Acronyms: LEDD, levodopa equivalent daily dose; MDS-UPDRS, Movement Disorder Society-Unified Parkinson’s Disease Rating Scale; OFF, medication “off” state. Significance codes: ***p < 0.001; **p < 0.01; *p < 0.05; †p ≈ 0.06. Responsiveness and dyskinesia omitted where non-significant; LEDD and duration shown only where significant. Outcomes are coded so that higher values indicate worse burden (more time in OFF or greater impact). Responsiveness is % OFF→ON improvement in MDS-UPDRS III; negative β therefore indicates that greater Responsiveness relates to less OFF burden.

| Group | Outcome | Across visits ($\beta$, p) | Between patients ($\beta$, p) |
| --- | --- | --- | --- |
| Pooled | OFF time (4.3) | MDS-UPDRS III OFF total; 0.12***; Freezing 0.06†; Tremor 0.07* | Responsiveness -0.13** |
|  | OFF impact (4.4) | MDS-UPDRS III OFF total; 0.13***; Freezing 0.09**; Dyskinesia 0.09** | Freezing 0.12**, Tremor 0.14***; Dyskinesia 0.10*, LEDD 0.14***; Duration 0.28*** |
| Early ($\leq 4$ y) | OFF time (4.3) | MDS-UPDRS III OFF total; 0.04; Tremor 0.12† | Responsiveness -0.33*** |
|  | OFF impact (4.4) | Tremor 0.12† | LEDD 0.38***; Tremor 0.20** |
| Late ($\geq 6$ y) | OFF time (4.3) | MDS-UPDRS III OFF total; 0.14***; Freezing 0.09*; Tremor 0.10** | Freezing 0.20**, Tremor 0.13* |
|  | OFF impact (4.4) | MDS-UPDRS III OFF total; 0.12**, Dyskinesia 0.08† | Freezing 0.24***; Tremor 0.13*; Dyskinesia 0.13*; Duration 0.29*** |

##### ≤4 years from diagnosis

For OFF time, within-person effects for motor, freezing, and tremor were small and non-significant; dyskinesia showed a trend (β = 0.093, p ≈ 0.060). Between patients, treatment responsiveness had the largest effect (β = −0.326, p < 0.0001). Sex also showed a positive association (β = 0.383, p = 0.014), indicating that being male was associated with greater OFF time in this group.

For OFF impact, within-person associations were minimal, with tremor showing a weak trend (β = 0.115, p ≈ 0.074). Between patients, LEDD showed the strongest association (β = 0.379, p < 1e−8), followed by tremor (β = 0.198, p = 0.005).

##### ≥6 years from diagnosis

For OFF time, within-person motor severity (β = 0.140, p < 0.001), freezing (β = 0.094, p = 0.034), and tremor (β = 0.100, p = 0.009) were associated with longer OFF periods. Between patients, freezing (β = 0.204, p = 0.004) and tremor (β = 0.132, p = 0.042) also showed positive associations, whilst the association of treatment responsiveness was negligible.

For OFF impact, within-person associations were modest (motor β = 0.124, p = 0.003; dyskinesia β = 0.079, p = 0.066). Between patients, disease duration (β = 0.289, p < 1e−8) and freezing (β = 0.198, p = 0.001) showed the strongest associations, with tremor (β = 0.133, p = 0.019) and dyskinesia (β = 0.134, p = 0.015) contributing smaller positive associations.

### Feature Importance (ΔR²)

We examined how much each predictor contributed to explaining OFF time and OFF impact. Rankings were based on two panels:

(1) Motor features (motor severity, freezing, tremor, treatment responsiveness, dyskinesia)
(2) Motor features plus covariates (LEDD and demographics such as sex, age at onset, and disease duration).

(See Supplementary Table S4 for full rankings.)

#### Pooled analyses

For OFF time, freezing (ΔR² = 1.57%) and motor severity (1.40%) were associated with the greatest increase in model fit among motor features, followed by treatment responsiveness (1.01%). Adding covariates did not change the order; demographic and LEDD effects added little (See Figure 1).

**Figure 1.**
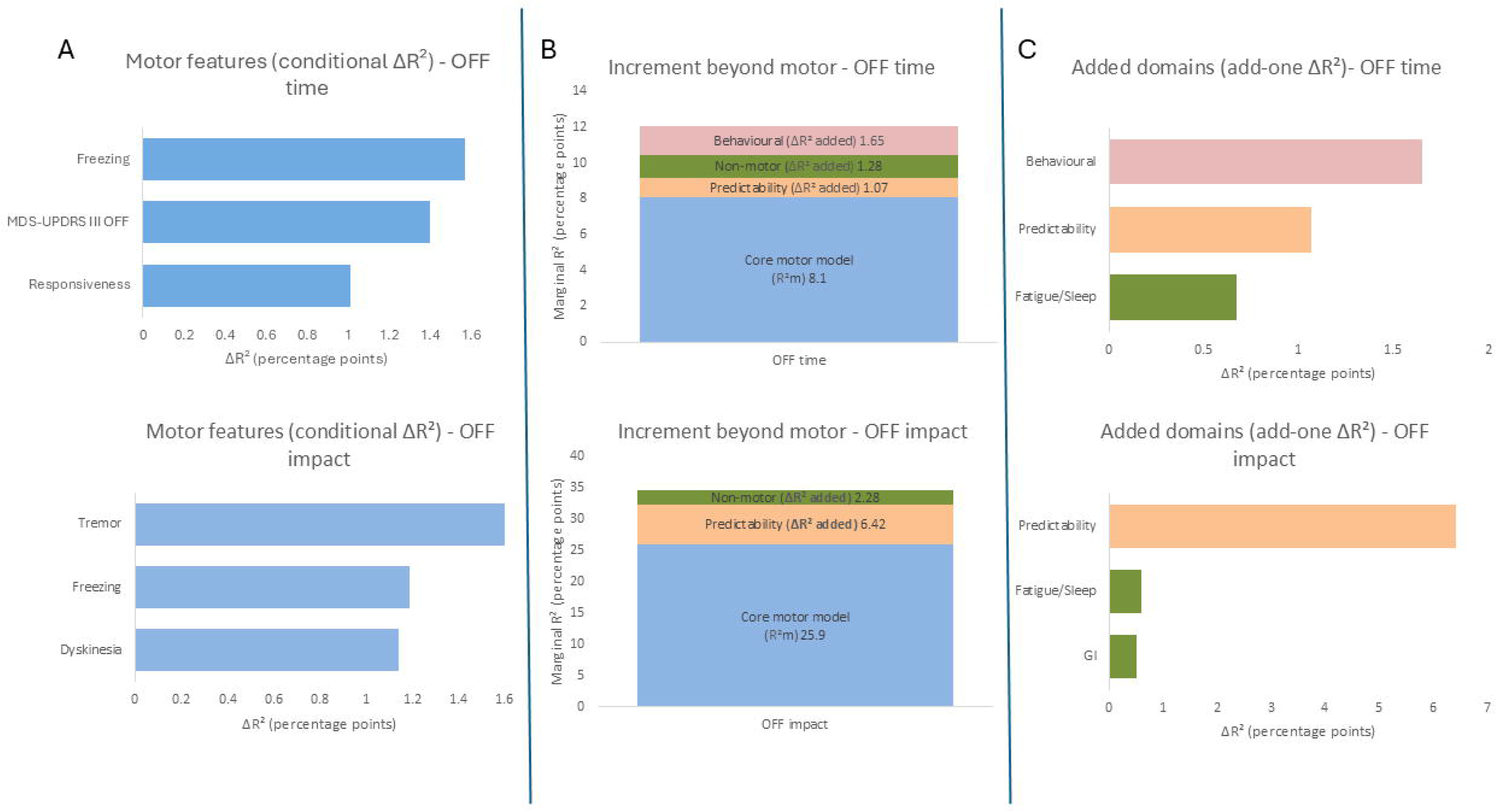
OFF burden model ladder: core motor model (with covariates) → increments from added domains → domain ranking. The three panels summarise how the core motor model and added domains relate to OFF burden. In this study, the core motor model denotes the prespecified motor feature set (MDS-UPDRS III OFF total; patient-reported freezing [II-2.13] and tremor [II-2.10]; levodopa responsiveness; dyskinesia burden) together with covariates (sex, age at onset, disease duration, person-mean LEDD). **Left (A): Motor features.** Within the motor feature set, ΔR² denotes the change in marginal R² attributable to a single feature, conditional on the other motor features (i.e., the loss when the feature is omitted; equivalently, the gain when it is added while other motor features are retained), ranking contributions for OFF time (IV 4.3) and OFF impact (IV 4.4). **Middle (B): Baseline and increments to combined R².** Bars show marginal R² (R²m) for the core motor model baseline, with the increment from added domain blocks stacked to the combined R²m. For presentation, the Non-motor block (Anxiety, Depression, Fatigue/Sleep, Autonomic, GI) is entered jointly. Because blocks overlap, ΔR² values are not unique or additive; the stacks are a communicative allocation of the combined R²m, not a strict variance partition. **Right (C): Added-domain ranking.** Among the added domains, ΔR² denotes the increase in R²m when the domain block is added on top of the core motor model baseline (add-one), ranking the strongest contributors for each outcome (e.g., Predictability for OFF impact). **Abbreviations.** ΔR², change in marginal R²; R²m, marginal R²; R²c, conditional R²; LEDD, levodopa equivalent daily dose; ESS, Epworth Sleepiness Scale; GDS-15, Geriatric Depression Scale-15; SCOPA-AUT, Scales for Outcomes in PD–Autonomic; QUIP/QUIP-RS, Questionnaire for Impulsive-Compulsive Disorders in PD; Predictability, MDS-UPDRS IV item 4.5

For OFF impact, tremor (1.60%), freezing (1.19%), and dyskinesia (1.14%) were associated with the largest increases among motor features. When covariates were included, disease duration (4.25%) and LEDD (1.68%) produced the greatest improvement in model fit, ahead of tremor (1.60%).

#### ≤4 years from diagnosis

For OFF time, responsiveness was the most influential feature (ΔR² = 7.23%), far exceeding motor severity (1.35%) and dyskinesia (0.74%). In the covariate-adjusted panel, responsiveness remained top-ranked, while sex (2.93%) displaced motor severity. For OFF impact, tremor (4.27%) led symptom contributions, followed by motor severity (1.42%) and freezing (1.37%). With covariates, LEDD explained the largest share (14.46%), ahead of tremor (4.27%) and motor severity (1.42%).

#### ≥6 years from diagnosis

For OFF time, freezing (2.80%) and tremor (2.03%) were the most prominent contributors; motor severity contributed modestly (1.29%). Age at onset (1.49%) entered the top three in covariate-adjusted models.

For OFF impact, freezing (2.20%), dyskinesia (1.74%), and tremor (1.43%) were the highest-ranked motor features. Disease duration contributed the largest share (5.24%) when covariates were included.

### Ordinal Sensitivity Analysis (CLMM)

Ordinal models produced patterns consistent with the linear mixed-effects models. For OFF time, motor severity and freezing remained the strongest correlates, with tremor contributing at both visit and patient levels. Treatment responsiveness and LEDD showed minimal associations. For OFF impact, disease duration and freezing (between-person) explained the largest share of ordinal variation, with additional contributions from motor severity and tremor. LEDD also contributed a small but statistically significant effect.

## Extended Analysis: Non-Motor, Temporal, and Behavioural Domains

### Individual addition of domains

Each prespecified domain, anxiety, depression, fatigue/sleep, autonomic and gastrointestinal symptoms, predictability (temporal), and impulse control behaviours (behavioural), was added separately to the core motor model. This approach allowed us to evaluate the association of each domain individually rather than grouping them together.

In pooled analyses, changes in marginal R² for OFF time were modest, with the largest contributions from impulse control behaviours (QUIP; ΔR² = +1.65%) and predictability (MDS-UPDRS IV item 4.5; +1.07%), followed by fatigue/sleep (ESS; +0.67%). For OFF impact, predictability contributed the largest share (+6.42%), with smaller increments from fatigue/sleep (+0.59%) and gastrointestinal symptoms (+0.50%).

In the ≤4-year group, predictability (+2.41%) and affective domains (anxiety +1.43%, depression +0.67%) contributed most to OFF time, while OFF impact was associated with predictability (+5.54%) and affective domains (anxiety +2.25%, depression +1.14%). See Figure 2.

**Figure 2.**
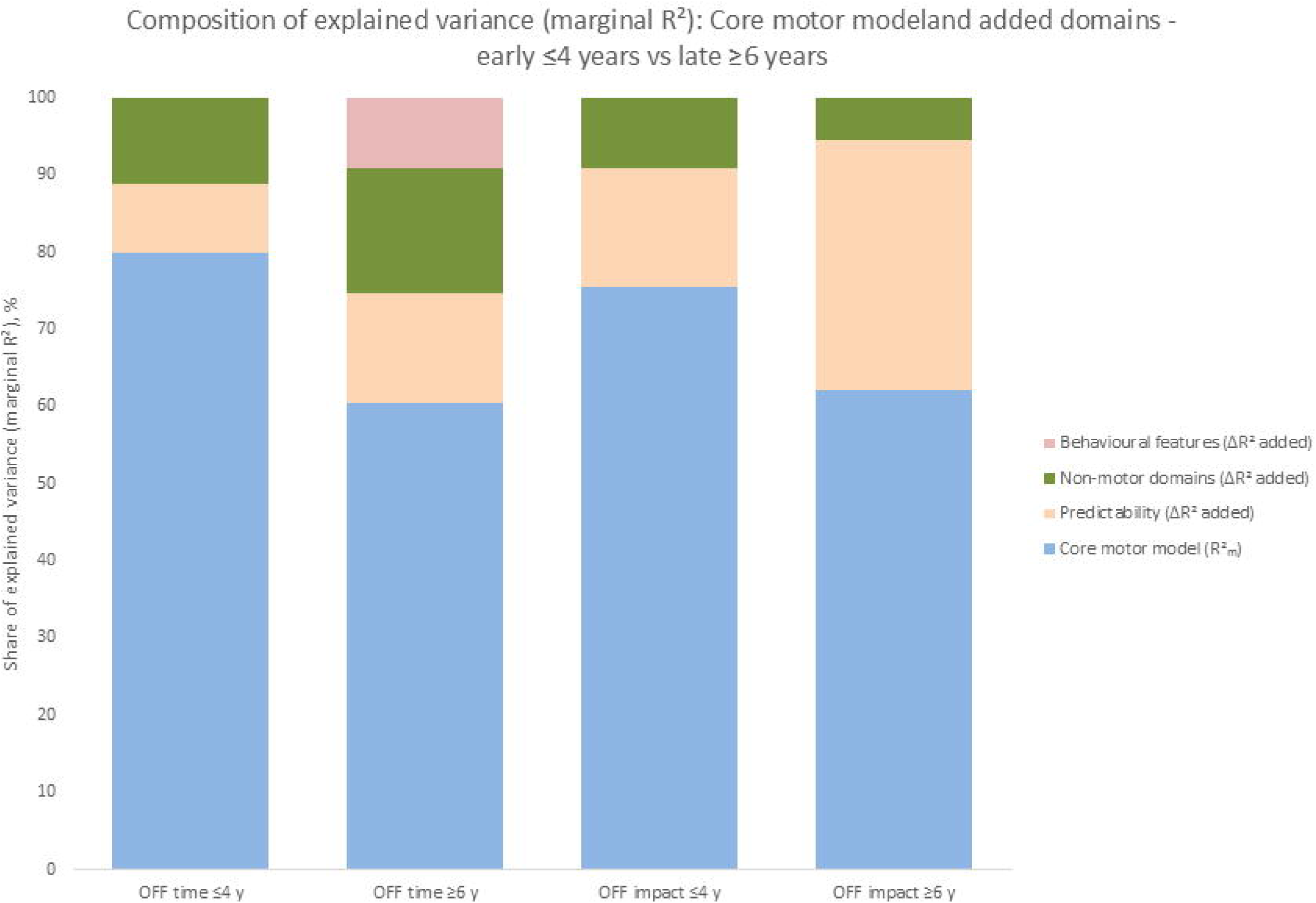
Composition of explained variance (marginal R²): core motor model baseline and increments from added domains. Each column displays the share of explained variance (R²m) for OFF time (IV 4.3) and OFF impact (IV 4.4), shown pooled and duration-stratified (Early ≤ 4 y; Late ≥ 6 y; visit-based). The lower segment is the core motor model baseline (MDS-UPDRS III OFF total; freezing; tremor; responsiveness; dyskinesia plus covariates: sex, age at onset, disease duration, person-mean LEDD). The upper segments are the increments from added domain blocks—Predictability (IV 4.5), Non-motor (Anxiety, Depression, Fatigue/Sleep, Autonomic, GI, entered jointly for presentation), and Behavioural (impulse-control behaviours). **Modelling note.** For domain blocks, ΔR² is the increase in R²m when the block is added on top of the core motor model baseline (add-one). Because blocks overlap, ΔR² values are non-additive; stacks provide an interpretable allocation of variance gained beyond the baseline, not a unique decomposition. **Abbreviations.** ΔR², change in marginal R²; R²m, marginal R²; LEDD, levodopa equivalent daily dose; ESS, Epworth Sleepiness Scale; GDS-15, Geriatric Depression Scale-15; SCOPA-AUT, Scales for Outcomes in PD–Autonomic; QUIP/QUIP-RS, Questionnaire for Impulsive-Compulsive Disorders in PD; Predictability, MDS-UPDRS IV item 4.5.

In the ≥6-year group, predictability (+2.64%) and impulse control behaviours (+1.72%) contributed most to OFF time, whereas OFF impact showed the largest association with predictability (+10.83%), with anxiety and depression contributing smaller shares. (See Table 4 for individual ΔR² rankings.)

**Table 4:**
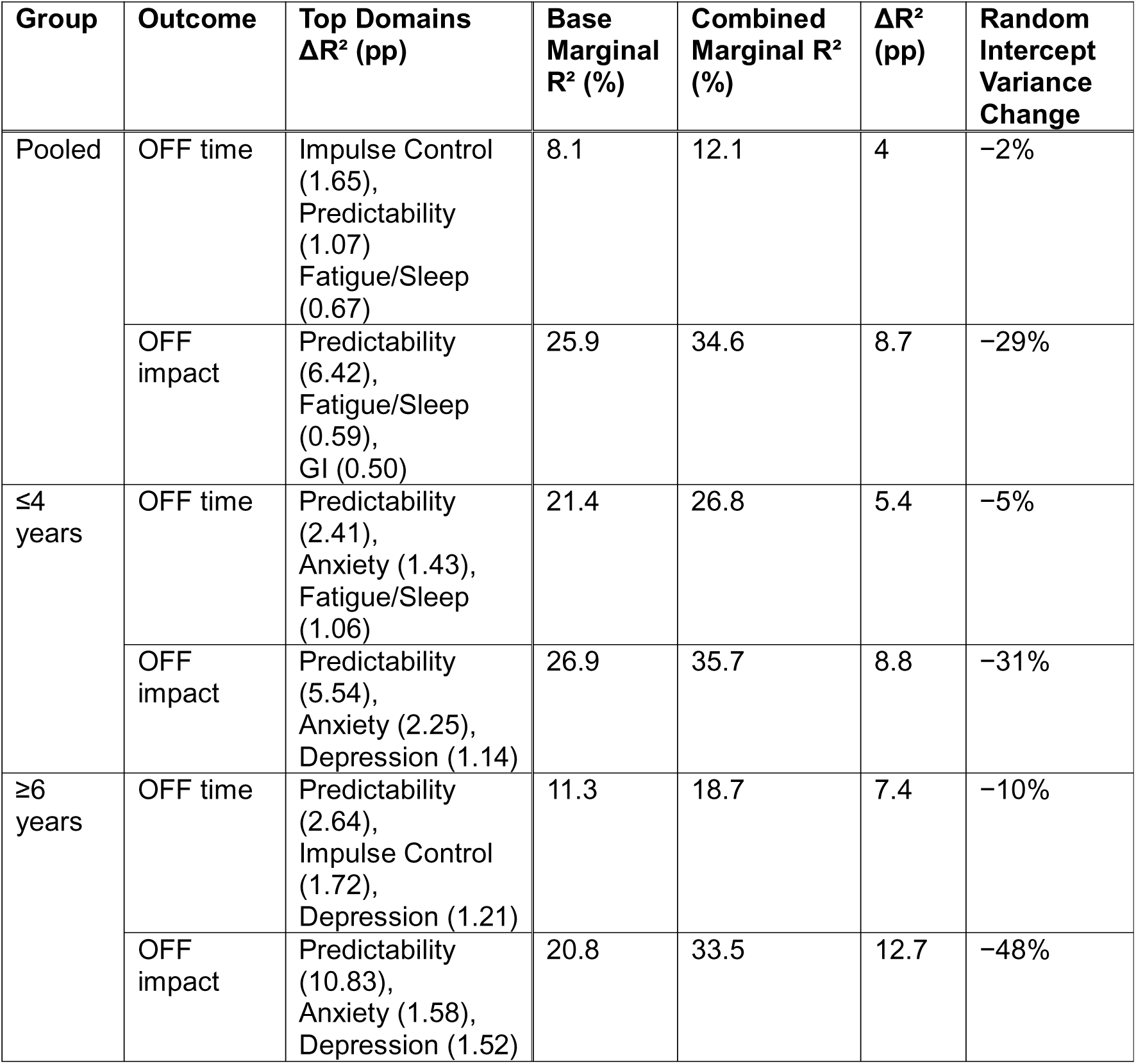
Summary of domain contributions and combined model fit for OFF time and OFF impact. Base marginal R² refers to the proportion of variance explained by fixed effects in the core motor model. Combined marginal R² reflects the variance explained after adding all prespecified extended domains (non-motor, temporal predictability, behavioural). ΔR² (pp) indicates the absolute increase in marginal R² in percentage points. Random Intercept Variance Change reflects the reduction in unexplained between-patient variability after adding extended domains. Top Domains (ΔR² %) lists the most influential individual domains when added separately to the motor model, based on their contribution to marginal R². Domains include non-motor symptoms (e.g., anxiety, depression, fatigue), temporal characteristics (predictability), and behavioural complications (impulse-control behaviours). Acronyms: R², coefficient of determination; pp, percentage points.

### Combined model (all domains)

When we added the non-motor, temporal, and behavioural domains to the core motor model, the fixed-effect contribution, representing how measured clinical features are associated with OFF burden, increased substantially for OFF impact (marginal R² +8.7 pp pooled; +12.7 pp in the ≥6-year group). At the same time, the random-effect variance, reflecting patient-to-patient differences not explained by these features, fell by up to ∼48%. This means that adding these domains helps account for why patients differ in perceived OFF impact beyond motor severity. For OFF time, the added value was modest (+4.0 pp pooled; +7.4 pp in the ≥6-year group), indicating OFF time remains more closely linked to motor severity.

(See Table 4)

### Mismatch residual analysis

Residual models assessed whether non-motor, temporal, and behavioural domains were associated with variation in OFF burden beyond motor severity. For OFF impact, predictability showed the largest association (ΔR² up to 13.5% in the ≥6-year group), with anxiety and depression contributing smaller shares. For OFF time, predictability and impulse control behaviours were the most prominent contributors in advanced disease. (See Supplementary Table S5 for mismatch residual results.)

### Sensitivity analyses

We repeated the models while adjusting for both OFF outcomes (e.g., OFF impact after accounting for OFF time). This did not change the ranking of associations.

(See Supplementary Table S6.)

Ordinal models produced patterns consistent with the main analysis. For OFF impact, both within-person and between-person predictability were associated with higher odds of reporting greater impact (OR 1.78 and OR 2.01 per SD, respectively), alongside associations for motor severity, freezing, dyskinesia, and disease duration. For OFF time, predictability also showed independent associations (OR 1.38 within; OR 1.86 between), with additional associations for depression (OR 2.33) and impulse control behaviours (negative association; OR 0.44).

(See Supplementary Table S7)

## Discussion

The core motor analysis revealed distinct association profiles for OFF time and OFF impact. OFF impact was more consistently explained by the included features, with fixed effects accounting for a larger share of variance than for OFF time. OFF time was most closely related to OFF-state motor severity and freezing, and in early disease it was lower among patients with stronger levodopa responsiveness. In contrast, OFF impact was more closely linked to disease duration and LEDD, suggesting that perceived burden reflects both symptom severity and long-term treatment exposure.

One explanation for this difference may be that patients find it easier to judge functional impairment than to estimate total OFF time. Prior studies have shown that OFF time reporting is vulnerable to recall bias and misclassification, especially when non-motor symptoms overlap(13, 14). In PD, classic experiments demonstrate underestimation of durations, with partial levodopa correction, consistent with a slowed “internal clock”(15).

Subsequent work confirms temporal-processing abnormalities, reinforcing caution when interpreting self-reported OFF durations(16, 17). By contrast, OFF impact is activity-anchored and likely judged more reliably.

The extended analysis was designed to quantify which domains beyond motor features contribute most to OFF burden, testing predictability, non-motor symptoms, and behavioural features against the core motor baseline. OFF is best understood as a spectrum, ranging from predictable end-of-dose wearing-off to on–off fluctuations, early-morning akinesia, delayed ON and dose failures. This heterogeneity helps explain why pattern complexity/unpredictability is linked to greater perceived impact in later disease(1, 18). In added-domain analyses, predictability emerged as the strongest correlate of OFF impact, particularly in later disease. Importantly, our results do not imply that unpredictability itself makes episodes intrinsically more severe; rather, unpredictability likely signals a different OFF phenotype, on–off fluctuations, which are often more abrupt and disabling than predictable end-of-dose wearing-off. This aligns with clinical experience and patient reports that irregular or complex OFF episodes are more disruptive than their duration alone. By contrast, associations with OFF time were modest; impulse-control behaviours and predictability contributed most consistently in later disease. Adding non-motor, temporal, and behavioural domains improved model fit for OFF impact and reduced unexplained between-patient variability, suggesting these domains help account for differences in perceived burden not captured by motor features alone.

Mechanistically, several factors may contribute to unpredictability, including medication adherence and timing (not assessed here). Peripheral mechanisms such as gastrointestinal dysfunction may impair levodopa absorption and contribute to delayed ON or dose failures, but GI symptom scores were not strongly associated with OFF burden, suggesting that peripheral factors alone are insufficient to explain the association(19, 20). Central mechanisms, including progressive loss of presynaptic dopamine storage and altered basal ganglia circuitry, may underlie more disruptive OFF episodes in later disease(1, 21–23).

Psychological factors such as fear and uncertainty may further amplify the impact of unpredictable fluctuations, consistent with qualitative reports that unpredictability undermines an individual’s planning and autonomy(6, 7). Taken together, these findings suggest that predictability reflects both central disease processes and the patient’s lived experience of uncertainty.

Separating within- from between-patient effects added clinical nuance. Within patients, higher OFF-state motor severity and lower predictability co-occurred with greater burden at a given visit, suggesting that management may be improved through same-day adjustments (timing/formulation, rescue strategies). Between patients, features reflecting more severe disease/ prolonged treatment exposure, including disease duration, mean LEDD and freezing, were more prominent associations for OFF impact, especially in ≥6 years, potentially suggesting that pathway-level decisions may be more helpful (e.g., earlier consideration of device-aided therapies).

To complement the domain-wise ΔR² feature analyses, we examined mismatch residuals, defined as the portion of OFF burden not explained by motor severity and covariates. This approach helps identify which features contribute to perceived burden among patients with similar motor profiles. Predictability remained the strongest correlate of OFF impact, particularly in later disease, with additional contributions from anxiety and depression. For OFF time, impulse-control behaviours and predictability were more prominent in the ≥6-year group. These findings suggest that the extended domains included in these analyses help explain why some patients report greater burden than expected based on motor features alone.

This study used data from the PPMI cohort, which recruited individuals with de novo PD. As a result, participants were younger and less advanced than many patients seen in routine clinical practice, particularly those being considered for device-aided therapies(9). The weighting of associations may differ in more advanced populations with severe motor fluctuations.

MDS-UPDRS Part IV items 4.3 (OFF time) and 4.4 (OFF impact) were analysed as continuous variables despite being ordinal (0–4). Sensitivity analyses using ordinal models produced similar rankings, supporting the validity of the main findings.

The dataset did not include detailed records of medication timing or adherence, meal timing, or non-motor fluctuation diaries, limiting our ability to explore the mechanisms underlying unpredictable OFF episodes. Non-motor symptoms were assessed globally rather than during OFF periods specifically, consistent with the use of patient-reported freezing and tremor (Part II) which capture functional burden over the preceding week rather than a state-specific snapshot. Evidence that non-motor symptoms worsen during OFF states(24, 25), and that global non-motor burden is associated with motor complication severity(26), supports the clinical relevance of these measures, though future studies incorporating OFF-specific non-motor assessment may refine these associations.

Finally, ΔR² values are model-dependent and tend to be conservative when predictors overlap. We report them within each outcome and disease-duration group to avoid cross-model comparisons and interpret them as relative contributions rather than absolute effect sizes.

These findings highlight the importance of assessing OFF period predictability in routine care. OFF impact appears closely linked to how reliably patients can anticipate and manage their symptoms, which has direct implications for treatment planning. Predictable end-of-dose wearing-off can often be managed by optimising baseline therapy (e.g., adjusting dose timing, using extended-release formulations, or adding COMT or MAO-B inhibitors). In contrast, OFF episodes that occur abruptly or irregularly, commonly referred to as on–off fluctuations, may additionally benefit from on-demand rescue therapies such as subcutaneous apomorphine, sublingual apomorphine, or inhaled levodopa(27–30). These agents provide rapid relief but may be underused due to availability or because OFF patterns are not systematically characterised in clinic.

Future research should test whether targeting predictability, through dose-timing aids, meal-timing counselling, long-acting or continuous dopaminergic strategies, adherence support, or rescue therapies, can reduce OFF impact. Studies incorporating timed medication and meal records, non-motor fluctuation tools, and objective monitoring are needed to clarify mechanisms and identify which patients benefit most. In parallel, objective wearables can complement patient-reported measures: inertial-sensor models already classify ON/OFF with high accuracy, and clinical deployments can detect unpredictable OFF and dyskinesia better than diaries; combining heart-rate variability with accelerometery may help forecast OFF-risk windows and guide dose-timing or rescue therapy(31).

Extending this work to advanced and device-treated cohorts will be essential to understand how the relative contribution of different domains shifts later in the disease and to integrate patient-reported triggers and coping strategies into care pathways.

Our findings also support moving beyond a binary “OFF vs not OFF” concept toward an axes-based model of OFF burden. Key axes include motor impairment and phenotype (e.g., OFF-state UPDRS III, freezing, tremor), levodopa responsiveness, temporal dynamics (predictability), affective state (anxiety, depression), fatigue, and autonomic and gastrointestinal symptoms. These axes contribute to OFF burden to varying degrees across the disease course. Early disease is dominated by predictable wearing-off, where motor severity and responsiveness matter most. Later disease brings greater unpredictability, in the form of ‘on-off fluctuations and non-motor or behavioural complications. A personalised toolkit approach, matching interventions to the dominant axes for each patient, may offer a practical way to individualise care. This framework aligns with progress toward individualised and precision-medicine approaches, focusing care on the elements of disease experience that are most impactful for each patient.

## Acknowledgements

The authors would like to thank all participants and investigators involved in the Parkinson’s Progression Markers Initiative (PPMI) for their invaluable contributions to this research. We are also grateful to the Michael J. Fox Foundation and its funding partners for supporting the PPMI study.

This work was supported by the Clinical Ageing Research Unit at Newcastle University and the Newcastle upon Tyne Hospitals NHS Foundation Trust. We acknowledge the infrastructure and collaborative environment provided by both institutions, which made this research possible.

Data used in the preparation of this article were based on the PPMI curated dataset dated 21^st^ March 2025, available from the Parkinson’s Progression Markers Initiative (PPMI) database (https://www.ppmi-info.org/access-data-specimens/download-data), RRID:SCR_006431. For up-to-date information on the study, visit http://www.ppmi-info.org.

PPMI – a public-private partnership – is funded by the Michael J. Fox Foundation for Parkinson’s Research and funding partners, including AbbVie, Alamar Biosciences, Aligning Science Across Parkinson’s (ASAP), Arrowhead Pharma, Arvinas, AskBio, BIAL, BioArctic, Biohaven, BlueRock Therapeutics, Bristol Myers Squibb, Calico Labs, Capsida Biotherapeutics, Critical Path Institute, DaCapo Brainscience, Denali, Edmond J. Safra Foundation, Eli Lilly, Gain Therapeutics, GE Healthcare, Genentech, GSK, Insitro, Johnson & Johnson Innovative Medicine, Lundbeck, Merck, Neumora, Neuron23, Novarti, Olink, Regeneron, Roche, Sanofi, Tenvie, UCB, Vanqua Bio, Voyager Therapeutics, The Weston Family Foundation.

## Author contributions

DL, MB, and NP conceptualised the investigation. DL, MB and NP, developed the methodology. DL performed formal analysis of the data. SS, RI, and CS validated the analysis. DL, CS, SS, VF, RI, DG, and NP contributed to conducting the investigation and collecting data. DL and NP prepared the original draft document. All authors contributed to the review of draft documents. All authors read and approved the final manuscript.

## Statements and Declarations

## Ethical Considerations

The Parkinson’s Progression Markers Initiative (PPMI) 2.0 study was approved by the London - City & East Research Ethics Committee (Ethics Code: 20/LO/0900) on September 7, 2020. All participating sites obtained ethical approval. This research was conducted ethically in accordance with the World Medical Association Declaration of Helsinki. The study is registered on ClinicalTrials.gov (Identifier: NCT01141023).

## Consent to Participate

All participants provided written informed consent prior to enrolment in the study.

## Declaration of conflicting Interest

The authors declared no potential conflicts of interest with respect to the research, authorship and/or publication of this article.

## Data availability

Data used in the preparation of this article was obtained on 2025-08-17 from the Parkinson’s Progression Markers Initiative (PPMI) database (www.ppmi-info.org/access-data-specimens/download-data), RRID:SCR_006431. For up-to-date information on the study, visit www.ppmi-info.org.

Protocol information for The Parkinson’s Progression Markers Initiative (PPMI) Clinical - Establishing a Deeply Phenotyped PD Cohort can be found at https://dx.doi.org/10.17504/protocols.io.n92ldmw6ol5b/v2. This study utilized PPMI Tier 1 clinical data.

Statistical analysis codes used to perform the linear mixed-effects modelling and mismatch residual analyses in this article are shared on Zenodo at https://doi.org/10.5281/zenodo.19421938.

## Notes

### Competing Interest Statement

The authors have declared no competing interest.

### Clinical Protocols

https://dx.doi.org/10.17504/protocols.io.n92ldmw6ol5b/v2

### Funding Statement

This study did not receive any funding

### Author Declarations

We used longitudinal data from the PPMI study, a multicentre observational study designed to identify PD progression markers (ClinicalTrials.gov: NCT01141023). All participants provided informed consent, and ethics approval was obtained at each site. The Parkinsons Progression Markers Initiative (PPMI) 2.0 study was approved by the London - City & East Research Ethics Committee (Ethics Code: 20/LO/0900) on September 7, 2020. All participating sites obtained ethical approval. This research was conducted ethically in accordance with the World Medical Association Declaration of Helsinki.

