## Supplementary Tables for "Beyond Motor Fluctuations: Understanding the Clinical Correlates of OFF burden in Parkinson’s Disease"

Supplementary Table 1A. Motor predictors and covariates: mechanistic justification, PPMI mapping, and rationale

| **Category** | **Mechanistic / clinical justification** | **PPMI variable(s) / derivation** | **Justification for chosen measure** | **Key refs** |
| --- | --- | --- | --- | --- |
| **Motor Predictors** | | | | |
| Motor severity | OFF‑state motor impairment reflects nigrostriatal denervation and anchors visit‑level motor burden. | MDS‑UPDRS Part III OFF total | Gold‑standard motor exam; OFF condition maximizes sensitivity to fluctuations. | Chahine 2020 (6),  Kelly 2019(4),  Sun 2020(28) |
| Freezing | Freezing contributes disproportionately to perceived disability | MDS‑UPDRS Part II item 2.13 (patient‑reported) | Patient report better captures between‑visit freezing and functional impact. | Chahine 2020(6),  Mantri 2021(7) |
| Tremor (functional impact) | Functional impact of tremor (buttons, writing) is not fully captured by Part III examination. | MDS‑UPDRS Part II item 2.10 (patient-reported) | Patient‑reported tremor reflects lived disability between visits. | Chahine 2020(6),  Mantri 2021(7) |
| Levodopa responsiveness | Percentage OFF→ON change. modulates OFF burden independent of baseline motor severity. | % Improvement in MDS UPDRS Part III OFF→ON (paired exam, same visit) | Clinically interpretable anchor used in PD trials and captures difference with ON state. | Kelly 2019(4) |
| Dyskinesia burden | Exposure‑related complications (time and impact) interact with OFF and treatment decisions. | MDS UPDRS Part IV 4.1 (time) + 4.2 (impact); composite = mean | Standard, validated items for dyskinesia in PD trials. | Chahine 2020(6) |
| **Covariates** | | | | |
| Demographics - Sex; Age at onset; Disease duration | Adjust for confounding and disease duration effects across outcomes. | Demographics; Disease duration from diagnosis (years) | Conventional covariate set for PD progression modelling. | Olanow 2013(2),  Kelly 2019(4),  Sun 2020(28),  Devraj 2024(29) |
| LEDD (exposure) | Dose both reflects severity and can contribute to complications via pulsatile stimulation; central and peripheral mechanisms. | LEDD, person‑mean across visits | Treating LEDD as an exposure covariate reduces bidirectional bias while retaining face validity. | Olanow 2013(2),  Kelly 2019(4),  Sun 2020(28) |

Supplementary Table 1B. Non‑motor and other predictors: mechanistic justification, PPMI mapping, and rationale

| **Category** | **Mechanistic / clinical justification** | **PPMI variable(s) / derivation** | **Justification for chosen measure** | **Key refs** |
| --- | --- | --- | --- | --- |
| Anxiety | Anxiety is common, fluctuates with OFF, and amplifies perceived burden (state and trait contributions). | STAI‑State (within‑person), STAI‑Trait (between‑person) | Widely used in PPMI; clinometric evaluation in PD including modern IRT/Rasch. | Kelly 2019(4) |
| Depression | Depressive symptoms worsen quality of life and perceived disability independent of motor state. | GDS‑15 | Brief, validated screening scale with strong clinical uptake in PD cohorts. | Kelly 2019(4) |
| Fatigue / sleepiness | Daytime sleepiness and fatigue exacerbate OFF‑related functional limitations. | Epworth Sleepiness Scale (ESS) | Simple, validated PRO; common in PD sleep research and registries. | Chahine 2020(6), Mantri 2021(7) |
| Autonomic / sensory | Autonomic symptoms (urinary, thermoregulation, light‑headedness) often worsen in OFF and increase burden. | SCOPA‑AUT total (minus GI domain) | PD‑specific autonomic scale with independent validations across languages. | Kelly 2019(4), Chahine 2020(6) |
| GI absorption | GI dysmotility/protein competition contribute to delayed‑on and dose failure, distinct from central mechanisms. | SCOPA‑AUT GI domain | GI sub score specifically targets absorption‑relevant complaints in PD. | Kelly 2019(4) |
| Predictability / irregularity | Unpredictable motor response increases patient burden. | MDS‑UPDRS IV item 4.5 (complexity of fluctuations) | Explicitly defined in MDS‑UPDRS | Chahine 2020(6) |
| Impulse control behaviours (ICD/DDS) | ICD/DDS can amplify distress and destabilize regimens, modifying OFF experience beyond motor severity. | QUIP / QUIP‑RS | Validated screening/rating tools. | Warren 2017(5), Kelly 2019(4) |

**Supplementary Tables S1A and S1B. Prespecified predictors of OFF burden: mechanistic justification, PPMI mapping, and rationale.**

Supplementary tables 1A and 1B summarize the predictors included in the analysis of OFF burden, grouped into motor predictors, covariates, and non‑motor or additional predictors. Each row details the mechanistic or clinical justification for inclusion, the corresponding Parkinson’s Progression Markers Initiative (PPMI) variable(s) or derived measure, and the rationale for selecting that instrument. Acronyms are expanded in alphabetical order as follows: CRF, case report form; DDS, dopamine dysregulation syndrome; ESS, Epworth Sleepiness Scale; FOG, freezing of gait; GDS‑15, 15‑item Geriatric Depression Scale; GI, gastrointestinal; ICD, impulse control disorder; LEDD, levodopa equivalent daily dose; MDS‑UPDRS, Movement Disorder Society‑Unified Parkinson’s Disease Rating Scale; OFF, medication “off” state; ON, medication “on” state; PPMI, Parkinson’s Progression Markers Initiative; PRO, patient‑reported outcome; QUIP, Questionnaire for Impulsive‑Compulsive Disorders in Parkinson’s Disease; QUIP‑RS, QUIP‑Rating Scale; SCOPA‑AUT, Scales for Outcomes in Parkinson’s Disease–Autonomic Dysfunction; STAI, State‑Trait Anxiety Inventory.

Motor predictors include MDS‑UPDRS Part III OFF total as the anchor for motor severity, patient‑reported freezing (Part II item 2.13), and tremor (Part II item 2.10), which better capture between‑visit functional impact than clinic‑based observation. Levodopa responsiveness was indexed as the percentage improvement in Part III from OFF to ON when paired examinations were available. Dyskinesia burden was summarized as the mean of Part IV items 4.1 (time) and 4.2 (impact). Covariates comprised sex, age at onset, disease duration, and person‑mean LEDD to account for cumulative exposure while minimizing bidirectional bias.

Non‑motor predictors were selected based on evidence linking these domains to OFF burden and mapped to validated PPMI instruments: anxiety (STAI‑State and STAI‑Trait), depression (GDS‑15), fatigue and sleepiness (ESS), autonomic and sensory symptoms (SCOPA‑AUT total minus GI domain), gastrointestinal absorption (SCOPA‑AUT GI domain), predictability of fluctuations (MDS‑UPDRS Part IV item 4.5), and behavioural complications (QUIP or QUIP‑RS). Items 4.3 (time in OFF) and 4.4 (OFF disability) were not reused as predictors to avoid endpoint contamination. Where repeated measures existed, within‑person and between‑person effects (e.g., STAI‑State vs STAI‑Trait; LEDD person‑mean) were modelled explicitly.

| **Variable** | **Group** | **N** | **Mean** | **SD** | **Median [Q1–Q3]** | **Min–Max** |
| --- | --- | --- | --- | --- | --- | --- |
| STAI State | Pooled | 1236 | 32.95 | 10.35 | 31 [24–39] | 20–75 |
|  | Early (≤4y) | 262 | 31.84 | 10.18 | 29 [24–38] | 20–70 |
|  | Late (≥6y) | 749 | 33.48 | 10.35 | 32 [25–40] | 20–75 |
| STAI Trait | Pooled | 1237 | 33.69 | 10.14 | 32 [25–40] | 20–75 |
|  | Early (≤4y) | 262 | 33.04 | 10.04 | 31 [25–39] | 20–64 |
|  | Late (≥6y) | 749 | 33.95 | 10.1 | 33 [25–40] | 20–75 |
| GDS | Pooled | 1236 | 3.03 | 3.01 | 2 [1–4] | 0–15 |
|  | Early (≤4y) | 260 | 2.77 | 2.9 | 2 [1–4] | 0–15 |
|  | Late (≥6y) | 749 | 3.11 | 3.01 | 2 [1–4] | 0–15 |
| ESS | Pooled | 1237 | 8 | 4.65 | 7 [5–11] | 0–24 |
|  | Early (≤4y) | 262 | 6.99 | 4.13 | 6 [4–9] | 0–21 |
|  | Late (≥6y) | 748 | 8.43 | 4.84 | 8 [5–11] | 0–24 |
| SCOPA AUT (total minus GI) | Pooled | 1231 | 10.91 | 6.01 | 10 [7–14] | 0–38 |
|  | Early (≤4y) | 260 | 9.83 | 5.85 | 9 [6–12] | 0–33 |
|  | Late (≥6y) | 744 | 11.44 | 6.06 | 11 [7–15] | 0–38 |
| SCOPA AUT (GI subscore) | Pooled | 1236 | 4.04 | 2.69 | 4 [2–6] | 0–18 |
|  | Early (≤4y) | 261 | 3.54 | 2.63 | 3 [2–5] | 0–18 |
|  | Late (≥6y) | 748 | 4.31 | 2.73 | 4 [2–6] | 0–18 |
| Predictability (UPDRS IV 4.5) | Pooled | 1252 | 1.22 | 0.83 | 1 [1–1] | 0–4 |
|  | Early (≤4y) | 265 | 1.12 | 0.89 | 1 [1–1] | 0–4 |
|  | Late (≥6y) | 760 | 1.28 | 0.83 | 1 [1–1] | 0–4 |
| QUIP | Pooled | 1239 | 0.38 | 0.8 | 0 [0–1] | 0–5 |
|  | Early (≤4y) | 261 | 0.34 | 0.71 | 0 [0–0] | 0–4 |
|  | Late (≥6y) | 751 | 0.41 | 0.83 | 0 [0–1] | 0–5 |

**Supplementary Table S2. Descriptive statistics for non-motor domains by group (pooled, early ≤4 years, late ≥6 years).**
Columns include N, mean, SD, median [Q1–Q3], and range (Min–Max). Acronyms: ESS, Epworth Sleepiness Scale; GDS, Geriatric Depression Scale; GI, gastrointestinal; QUIP, Questionnaire for Impulsive-Compulsive Disorders in Parkinson’s Disease; SCOPA-AUT, Scales for Outcomes in Parkinson’s Disease-Autonomic Dysfunction; STAI, State-Trait Anxiety Inventory; MDS-UPDRS, Movement Disorders Society Unified Parkinson’s Disease Rating Scale.

| **Level** | **Pooled** | **Early (≤4 y)** | **Late (≥6 y)** |
| --- | --- | --- | --- |
| 0 | 9.90% | 17.40% | 7.40% |
| 1 | 70.70% | 66.40% | 70.00% |
| 2 | 10.70% | 6.40% | 13.90% |
| 3 | 5.20% | 6.40% | 4.70% |
| 4 | 3.50% | 3.40% | 3.90% |

**Supplementary Table S3. Predictability of fluctuations (MDS-UPDRS Part IV item 4.5) distribution by group (pooled, early ≤4 years, late ≥6 years).**
Values represent the percentage of visits at each level (0–4). Acronyms: MDS-UPDRS, Movement Disorder Society-Unified Parkinson’s Disease Rating Scale.

| **Group** | **Outcome** | **Rank** | **Motor-only Feature** | **Motor-only ΔR² (%)** | **Motor + Covariates Feature** | **Motor + Covariates ΔR² (%)** |
| --- | --- | --- | --- | --- | --- | --- |
| Pooled | 4.3 – OFF time | 1 | Freezing | 1.57 | Freezing | 1.57 |
|  |  | 2 | OFF score | 1.4 | OFF score | 1.4 |
|  |  | 3 | Responsiveness | 1.01 | Responsiveness | 1.01 |
|  | 4.4 – OFF impact | 1 | Tremor | 1.6 | Duration | 4.25 |
|  |  | 2 | Freezing | 1.19 | LEDD | 1.68 |
|  |  | 3 | Dyskinesia | 1.14 | Tremor | 1.6 |
| Early (≤ 4 y) | 4.3 – OFF time | 1 | Responsiveness | 7.23 | Responsiveness | 7.23 |
|  |  | 2 | OFF score | 1.35 | Sex | 2.93 |
|  |  | 3 | Dyskinesia | 0.74 | OFF score | 1.35 |
|  | 4.4 – OFF impact | 1 | Tremor | 4.27 | LEDD | 14.46 |
|  |  | 2 | OFF score | 1.42 | Tremor | 4.27 |
|  |  | 3 | Freezing | 1.37 | OFF score | 1.42 |
| Late (≥ 6 y) | 4.3 – OFF time | 1 | Freezing | 2.8 | Freezing | 2.8 |
|  |  | 2 | Tremor | 2.03 | Tremor | 2.03 |
|  |  | 3 | OFF score | 1.29 | Age at onset | 1.49 |
|  | 4.4 – OFF impact | 1 | Freezing | 2.2 | Duration | 5.24 |
|  |  | 2 | Dyskinesia | 1.74 | Freezing | 2.2 |
|  |  | 3 | Tremor | 1.43 | Dyskinesia | 1.74 |

**Supplementary Table S4. Top contributors to variance explained (ΔR²) for OFF time and OFF impact across panels and groups.**
ΔR² represents the change in marginal R² when each construct is removed, quantifying its unique contribution to model fit. Rankings are shown for two panels: Motor-only (key motor symptom domains: clinician-assessed OFF score, patient-reported freezing, tremor, Responsiveness, and dyskinesia) and Motor + Covariates (adding LEDD and demographics: sex, age at onset, disease duration). Acronyms: LEDD, levodopa equivalent daily dose; OFF score, MDS-UPDRS Part III OFF total; Responsiveness, % improvement from OFF to ON in MDS-UPDRS Part III.

| **Group** | **Outcome** | **Axis** | **ΔR² (%)** |
| --- | --- | --- | --- |
| Pooled | OFF time (4.3) | Impulse Control Behaviours (QUIP) | 1.62 |
|  |  | Predictability | 1.18 |
|  | OFF impact (4.4) | Predictability | 8.48 |
|  |  | Fatigue/Sleep | 0.83 |
| Early (≤4 y) | OFF time (4.3) | Predictability | 2.41 |
|  |  | Anxiety | 1.43 |
|  | OFF impact (4.4) | Predictability | 5.54 |
|  |  | Anxiety | 2.25 |
| Late (≥6 y) | OFF time (4.3) | Predictability | 2.64 |
|  |  | Impulse Control Behaviours (QUIP) | 1.72 |
|  | OFF impact (4.4) | Predictability | 13.45 |
|  |  | Anxiety | 0.64 |

**Supplementary Table S5. Variance explained (ΔR²) by non‑motor axes in residual OFF burden after adjusting for the motor model.**
Values are the additional marginal R² explained by each axis in mismatch models; the top two axes per outcome and group are displayed (full axis rankings are provided in the Supplementary Materials). Note: ΔR² quantifies contribution to model fit and does not imply direction. In CLMM sensitivity analyses, most axes had positive associations with OFF burden; an exception was Impulse Control Behaviours (QUIP) for OFF time (negative between‑person association).

| Group | Conditioned outcome† | R²m | R²c |
| --- | --- | --- | --- |
| Pooled | 4.4 \| motor+4.3 | 0.1179 | 0.2815 |
|  | 4.3 \| motor+4.4 | 0.0338 | 0.4019 |
| Early (≤4 y) | 4.4 \| motor+4.3 | 0.1129 | 0.2631 |
|  | 4.3 \| motor+4.4 | 0.074 | 0.6013 |
| Late (≥6 y) | 4.4 \| motor+4.3 | 0.148 | 0.1915 |
|  | 4.3 \| motor+4.4 | 0.0458 | 0.3714 |

**Supplementary Table S6A.** **Model fit for conditioned residual analyses.** R²m = marginal R² (fixed effects), R²c = conditional R² (fixed + random effects). †“Conditioned outcome” indicates the target outcome modelled after adjusting for the motor block and the other OFF outcome (e.g., “4.4 | motor+4.3” models OFF impact beyond motor severity and OFF time).

| **Group** | **Conditioned outcome†** | **Axis** | **ΔR² (%)** |
| --- | --- | --- | --- |
| Pooled | 4.4 \| motor+4.3 | Predictability | 8.38 |
|  |  | Fatigue/Sleep | 0.64 |
|  |  | GI | 0.59 |
|  |  | Autonomic | 0.2 |
|  | 4.3 \| motor+4.4 | Impulse Control Behaviours (QUIP) | 1.67 |
|  |  | Predictability | 1.19 |
|  |  | Fatigue/Sleep | 0.46 |
|  |  | Anxiety | 0.18 |
|  |  | Depression | 0.15 |
| Early (≤4 y) | 4.4 \| motor+4.3 | Predictability | 5.23 |
|  |  | Anxiety | 2.08 |
|  |  | Depression | 1.04 |
|  |  | Fatigue/Sleep | 1.03 |
|  |  | GI | 0.63 |
|  | 4.3 \| motor+4.4 | Predictability | 2.25 |
|  |  | Anxiety | 1.34 |
|  |  | Fatigue/Sleep | 1.12 |
|  |  | GI | 0.65 |
|  |  | Depression | 0.63 |
| Late (≥6 y) | 4.4 \| motor+4.3 | Predictability | 11.3 |
|  |  | Anxiety | 0.79 |
|  |  | Impulse Control Behaviours (QUIP) | 0.55 |
|  |  | GI | 0.1 |
|  | 4.3 \| motor+4.4 | Impulse Control Behaviours (QUIP) | 2.23 |
|  |  | Depression | 1.34 |
|  |  | Anxiety | 0.8 |
|  |  | Predictability | 0.45 |

**Supplementary Table S6B. Axis-level ΔR² (%) for conditioned residual models.** Values show the additional marginal R² explained by each non‑motor axis after adjusting for the motor block and the alternate OFF outcome. ΔR² reflects contribution to fit, not direction of effect. In the CLMM sensitivity (late group), most axes showed positive associations with OFF burden; Impulse Control Behaviours (QUIP) showed a negative between‑person association with OFF time, plausibly reflecting higher dopaminergic exposure. Acronyms: QUIP, Questionnaire for Impulsive‑Compulsive Disorders in Parkinson’s Disease; GI, gastrointestinal.

| **Outcome** | **N** | **LogLik (full)** | **LogLik (null)** | **AIC** | **BIC** | **R² (Cox–Snell)** | **R² (Nagelkerke)** |
| --- | --- | --- | --- | --- | --- | --- | --- |
| 4.4 (OFF impact) | 475 | −551.358 | −646.631 | 1170.71 | 1312.26 | 0.3304 | 0.35369 |
| 4.3 (OFF time) | 475 | −250.345 | −296.085 | 566.69 | 704.08 | 0.1751 | 0.24585 |

**Supplementary Table S7A. Model fit for cumulative link mixed models (CLMM) in the ≥6 years group.**
The table reports the number of visits included (N), log-likelihood for the full and null models (LogLik), Akaike Information Criterion (AIC), Bayesian Information Criterion (BIC), and pseudo-R² values (Cox–Snell and Nagelkerke). The null model used for pseudo-R² comparisons included threshold parameters and a participant random intercept only. Higher pseudo-R² values indicate greater explanatory value on the ordinal (logit) scale. OFF impact refers to MDS-UPDRS Part IV item 4.4 (impact of OFF periods), and OFF time refers to item 4.3 (time spent in OFF).

| **Outcome** | **Predictor (standardized, per SD)** | **OR** | **95% CI** | **p-value** |
| --- | --- | --- | --- | --- |
| 4.4 – OFF impact | Predictability (within-person) | 1.78 | 1.46–2.17 | 1.50×10⁻⁸ |
|  | Predictability (between-person) | 2.01 | 1.55–2.60 | 1.27×10⁻⁷ |
|  | Disease duration (between-person) | 2.43 | 1.69–3.49 | 1.81×10⁻⁶ |
|  | MDS UPDRS‑III OFF (within-person) | 1.27 | 1.05–1.53 | 0.014 |
|  | Freezing (between-person) | 1.38 | 1.07–1.79 | 0.014 |
|  | Dyskinesia (between-person) | 1.25 | 1.01–1.54 | 0.041 |
|  | Age at onset (between-person) | 0.76 | 0.59–0.97 | 0.03 |
| 4.3 – OFF time | MDS UPDRS‑III OFF (within-person) | 1.73 | 1.25–2.39 | 8.63×10⁻⁴ |
|  | Predictability (between-person) | 1.86 | 1.18–2.93 | 0.0073 |
|  | Predictability (within-person) | 1.38 | 1.07–1.80 | 0.014 |
|  | Depression — GDS (between-person) | 2.33 | 1.17–4.64 | 0.016 |
|  | Tremor (within-person) | 1.32 | 1.00–1.75 | 0.0497 |
|  | Impulse Control Behaviours — QUIP (between-person) | 0.44 | 0.26–0.76 | 0.0031 |
|  | Age at onset (between-person) | 0.4 | 0.25–0.65 | 2.00×10⁻⁴ |

**Supplementary Table S7B.** **Odds ratios (OR) from cumulative link mixed models (CLMM) for significant predictors (p < 0.05) in the late group (≥6 years).**

The table reports predictors associated with OFF impact (MDS-UPDRS Part IV item 4.4) and OFF time (item 4.3). OR values represent the change in odds of being in a higher OFF category per one standard deviation increase in the predictor. “Within-person” refers to visit-level deviations from a participant’s mean, and “between-person” refers to the participant’s mean (trait effect). Confidence intervals (CI) are shown in brackets. Models included the full motor block, non-motor axes, and covariates with a random intercept for participant. Acronyms: GDS, Geriatric Depression Scale; QUIP, Questionnaire for Impulsive-Compulsive Disorders in Parkinson’s Disease; MDS-UPDRS, Movement Disorders Society Unified Parkinson’s Disease Rating Scale.
